# Circulating proteomic profiles are associated with the onset of type 2 diabetes in a multi-ethnic Asian population – a longitudinal study

**DOI:** 10.1101/2024.05.28.24308009

**Authors:** Yujian Liang, Charlie G.Y. Lim, Scott C. Ritchie, Nicolas Bertin, Jin-Fang Chai, Jiali Yao, Yun Li, E Shyong Tai, Rob M. van Dam, Xueling Sim

## Abstract

Type 2 diabetes (T2D) is a major global concern, with Asia at its epicenter in recent years. Proteins, products of gene transcription, serve as dynamic biomarkers for pinpointing perturbed pathways in disease development. To understand the protein function in incident T2D, we examined the association of 4,775 plasma proteins with incident T2D in a Singapore multi-ethnic cohort of 1,659 Asian participants (539 cases and 1,120 controls). Our analysis revealed 522 proteins that were associated with incident T2D after adjusting for age, sex, and ethnicity. Among the 522 proteins associated with incident T2D, the change in 205 plasma proteins, observed in parallel with the development of T2D at baseline and six-years follow-up, were further associated with incident T2D. The associated proteins showed enrichment in neuron generation, glycosaminoglycan binding, and insulin-like growth factor binding. Two-sample Mendelian randomization analysis suggested three plasma proteins, GSTA1, INHBC, and FGL1, play causal roles in the development of T2D, with colocalization evidence supporting GSTA1 and INHBC. Our findings reveal plasma protein profiles linked to the onset of T2D in Asian populations, offering insights into the biological mechanisms of T2D development.

**Article Highlights:**

- We measured 4,775 proteins in 1,659 Asian participants and reported 522 proteins (316 novel) associated with incident type 2 diabetes (T2D).
- Of 479 proteins previously reported to be associated with T2D, 384 (80.17%) showed same direction of effects in our study.
- Many of the incident T2D-associated proteins were also associated with insulin resistance markers including waist circumference and triglycerides.
- Mendelian randomization analysis identified three proteins, GSTA1, INHBC, and FGL1, to have plausible casual associations with T2D development. Colocalization analysis and previous studies further supported GSTA1 and INHBC as candidate causal proteins.

## Introduction

Type 2 diabetes (T2D) and its complications significantly contribute to the burden of mortality and disability globally [1, 2]. Lifestyle factors, genetic predisposition, and other risk factors jointly play a role in the pathogenesis of T2D, resulting in multiple dysfunctional pathways involving many organ systems [3, 4]. More than 650 T2D-associated genetic signals have been identified in trans-ancestry genetic association studies, but translating these genetic associations to better understand T2D pathophysiology has not been straightforward [5–9]. Most associated variants are in noncoding regions, and the molecular mechanisms linking variants to T2D development are unclear [6]. In addition, genetic variants fail to capture the effects of lifestyle exposures and their interactions with genetic factors leading to T2D.

Proteins, as products of gene transcription, function as dynamic biomarkers reflecting genetic regulation and environmental changes, which can aid in pinpointing the perturbed proteins and pathways in T2D pathogenesis [10]. Large-scale population proteomic studies on plasma proteins and complex diseases, including T2D, have become possible in recent years as high-throughput proteomic profiling technologies improve [11–16]. Using the genome, Mendelian randomization (MR) uses genetic variants to assess causality and has been used to integrate the genome and proteome to reveal causal proteins and potential T2D therapeutic targets [12, 13, 16, 17]. Many proteins, including ACY1, MXRA8, and ADIPOQ, are associated with incident T2D after accounting for factors such as body mass index (BMI) and fasting plasma glucose [12, 13], and highly consistent across European and African American populations [15]. In a recent study including 5,438 individuals from Iceland, 536 proteins were associated with prevalent and/or incident T2D, and 16 of these proteins had a potential causal link to T2D [12]. As the pathophysiology of T2D may vary across populations [18, 19], the transferability of these findings and novel proteins to Asian populations can highlight similar and unique pathways associated with the onset of T2D and to aid in the understanding of this disorder.

Using data from 1,659 participants (539 incident cases and 1,120 controls) of the Singapore Multi-Ethnic Cohort Phase 1 (MEC1), we examined the association of 4,775 plasma proteins with incident T2D. In a subset of participants, we further assess the change in plasma proteins with the development of T2D using repeated measurements collected at two timepoints (baseline and follow-up). Finally, we evaluated the causality of the relationship between these proteins and T2D using MR and colocalization analyses to identify potential therapeutic targets.

## Methods

### Study cohort

Participants in this study were sampled from the Singapore MEC1, a population-based cohort study that aims to discover how lifestyle factors, genetic factors, and their interactions impact the development of chronic health conditions in Singapore (https://blog.nus.edu.sg/sphs/population-studies/multi-ethnic-cohort-phase-1-mec1/) [20]. Participants were recruited between 2004 and 2010 for a baseline assessment and were invited to a follow-up visit between 2011 and 2016 (mean follow-up duration of 6.3 ± 1.6 years). At both baseline and follow-up, participants completed a standardized interviewer-administered questionnaire that included questions on socio-demographic and lifestyle factors, personal and family medical history, and medication use. Participants underwent a physical examination where anthropometric and blood pressure measurements were taken by trained staff (**Supplementary Note**) [20]. Fasting blood samples were drawn for biomarker measurements and stored at -80°C on the same day. Written consent was obtained from all participants, and this study was approved by the National University of Singapore Institutional Review Board (reference codes: B-16-158 and N-18-059).

Using MEC1, we carried out a nested case-control study for incident T2D. Incident T2D cases were ascertained through linkage with records on the national healthcare database, self-reported history of T2D at follow-up (physician diagnosis or use of diabetes medication), or having fasting glucose (≥7 mmol/L or hemoglobin A1c (HbA1c) (≥6.5%) levels at follow-up following the American Diabetes Association criteria for diabetes [21]. Controls were selected by incident density sampling matching (1 case to 2 controls) by age (±5 years), sex, ethnicity, and date of blood collection (±2 years). The final sample for the analysis of baseline proteomics and incident T2D association included 539 cases and 1,120 controls (**Supplementary Figure 1** and **Supplementary Note**). For protein change analysis, we only included newly diagnosed cases at follow-up using fasting glucose or HbA1c, resulting in 216 cases and 458 controls at baseline and follow-up (T2D incidence) (**Supplementary Figure 1** and **Supplementary Note**).

### Protein profiling

The relative concentrations of proteins were measured in plasma samples from MEC1 participants using the aptamer-based SomaScan assay v4 platform (SomaLogic Inc., Boulder, CO) [22] (https://mohanlab.bme.uh.edu/wp-content/uploads/2017/02/SSM-002-Rev-4-SOMAscan-Technical-White-Paper.pdf). SomaLogic employed an in-house library (N_aptamers_ = 5,272) of modified Slow Off-rate Modified Aptamers (SOMAmers) to target specific proteins. The protein concentrations were quantified using a DNA microarray and expressed in terms of relative fluorescent units (RFU).

The routine quality control processes used by SomaLogic consist of several steps with different control samples as the reference (**Supplementary Note**). After normalization, 82 samples were excluded with scale factors beyond the range of acceptance criteria (i.e., 0.4 to 2.5) (**Supplementary Figure 1**). In addition, we applied principal component analysis (PCA) on log_2_-transformed protein levels at the sample and aptamer level to identify potential outliers. No additional samples or aptamers were excluded due to excessive principal component values and/or interquartile ranges values (**Supplementary Figure 2** and **Supplementary Figure 3**). The median coefficient of variation (CV) for aptamers with both Calibrator Controls and Quality Controls were below 15% for all plates (**Supplementary Figure 4**). Based on the aptamer annotation, we excluded 294 aptamers, such as non-human proteins, non-protein aptamers, and aptamers without Entrez gene symbols or UniProt IDs. Finally, 4,978 aptamers targeting 4,775 unique proteins or protein isoforms remained for subsequent analysis.

### Statistical Analysis

We applied two transformations to protein RFU levels at each timepoint: log_2_ transformation for interpretability of the effect sizes and rank-based inverse normal transformation to calculate *P*-values to assess statistical significance. We reported the effect sizes obtained from log_2_-transformed protein level associations and their corresponding *P*-values resulting from rank-based inverse normal transformation, unless otherwise specified. The log_2_-transformed protein levels and phenotypic trait values at different time points were winsorized separately within a range of ±5 standard deviations (SD) to reduce the influence of outliers. Insulin and triglycerides (TG) levels were log-transformed due to skewed distributions. Logistic regression model was used to assess the association of baseline protein levels with incident T2D with adjustment for age, sex, and ethnicity (Model 1) to maximise samples that no longer had matched case/control due to sample loss (**Supplementary Note** and **Supplementary Figure 5**). A Bonferroni threshold of *P* = 1.05 × 10^-5^ (0.05/4,775 proteins) was considered significant. If more than one aptamer was targeted for the same protein, only the aptamer with the smallest *P* value in Model 1 was retained for subsequent analysis. We sequentially included fasting glucose (Model 2), waist circumference (Model 3), and TG (Model 4). The final model was derived using stepwise logistic regression analysis using the Bayesian information criterion (BIC), which imposes a higher penalty on each parameter and tends to select simpler models. Variables in the full model (Model 5) included age, sex, ethnicity, fasting glucose, waist circumference, TG, systolic blood pressure (SBP), and family history of diabetes. Based on the BIC, other T2D-associated variables, such as BMI, hip circumference, low-density lipoprotein (LDL), C-reactive protein, blood pressure-lowering medication, and a history of hypertension, were not selected for Model 5 (**Supplementary Table 1**).

For each of the plasma proteins associated with incident T2D in our study, we further examined the associations between their longitudinal changes over time with T2D. We first computed the log_2_-transformed fold change in protein levels between two time points, referred to as the ’protein difference’. Similar to the baseline association, we used this protein difference for effect size interpretability, and both the protein differences and the baseline protein levels were winsorized within a range of ±5 SD. Additionally, we applied a rank-based inverse normal transformation of these protein differences to assess the statistical significance. Logistic regression models were used to investigate the relationship between these protein differences and incident T2D. The basic model adjusted for age at baseline, sex, ethnicity, and corresponding baseline protein level. We sequentially included in baseline fasting glucose, family history of diabetes, and changes in waist circumference, TG, and SBP between baseline and follow-up into the models. All analyses were performed in R version 4.2.2.

### Functional enrichment analysis

Enrichment analyses of baseline proteins and changes in proteins associated with incident T2D were performed to identify biological pathways. We applied g:Profiler to perform functional enrichment for each protein category [23] with a significance threshold of Benjamini-Hochberg false discovery rate (FDR) corrected *P*-value less than 0.05. We also estimated tissue specificity using TissueEnrich with Human Protein Atlas data [24]. Both enrichment analyses used the SomaScan v4 panel protein (N = 4,775) as the background list (**Supplementary Note**).

### Whole-genome data

Whole genome data were available for a subset of MEC1 participants as National Precision Medicine Programme Phase I (https://npm.a-star.edu.sg/) [25]. Samples were whole-genome sequenced to an average of 15X coverage. Read alignment was performed with BWA-MEM (version 0.7.17). Variant discovery and genotyping were performed with GATK (version 4.0.6.0). Site-level filtering included only retaining VQSR-PASS and non-STAR allele variants. At the sample level, samples with call rates <95%, BAM cross-contamination rates >2%, or BAM error rates >1.5% were excluded. At the genotype call level, genotypes with depth (DP) <5 in any ancestry group or genotype quality (GQ) <20 or allele balance (AB) >0.8 (heterozygotes calls) were set to missing. Genetic variants were filtered to exclude those with robust, unified test for Hardy-Weinberg equilibrium (RUTH) *P*-value <0.01, a variant call rate <90%, being monomorphic, or having a minor allele count (MAC) <5 prior to phasing with Eagle version 2.4 [26, 27]. After quality control, the dataset included 16,014,113 genetic variants in a sample set of 1,753 individuals with both genetic and proteomic data.

### Mendelian randomization (MR)

To evaluate the potential causal role of the proteins associated with T2D, we performed two-sample MR analyses using multi-ancestry protein quantitative trait locus (pQTL) from our study and summary statistics from independent multi-ancestry genome-wide association studies (GWAS) for T2D (N = 1,339,889) (**Supplementary Note**) [9]. We identified genetic instruments of proteins by examining independent genetic variants within a 1Mb local region around each protein-encoding gene (**Supplementary Note**). The inverse variance-weighted (IVW) method was used to estimate the combined effect size of multiple instrumental variables, and the Wald ratio was used when only one variant instrument was available. FDR-corrected *P*-values less than 0.05 were considered significant. We used the Cochran Q statistic to assess instrument heterogeneity for proteins with more than one genetic instrument and the MR-Egger regression intercept to detect horizontal pleiotropy for proteins with more than two genetic instruments. A non-zero intercept in MR-Egger regression indicates the presence of pleiotropy in the variant [28]. R package “TwoSampleMR” was used for MR analyzes [29].

### Colocalization analysis

To assess the colocalization of the pQTLs and T2D association, we estimated the posterior probability of a shared genetic variant for both protein and T2D within a Bayesian framework at each locus [30]. Using the same independent multi-ancestry T2D GWAS summary statistics used for MR analysis [9], colocalization evidence was evaluated using posterior probabilities (PP) for hypothesis 4 (H4), indicating the joint presence of associations for both traits driven by the same causal variant. Associations with PP H4 > 0.5 were defined as ‘likely to colocalize’, while PP H4 > 0.8 indicated a ’highly likely’ colocalization. The colocalization analysis was performed utilizing the ‘coloc’ package in R.

## Results

### Study samples characteristics

Figure 1 summarizes the study design and the number of samples included for each analysis. For the association of baseline proteins and T2D incidence, we included 539 incident T2D cases (218 Chinese, 165 Malay, 156 Indian) and 1,120 controls (409 Chinese, 348 Malay, 363 Indian). For the longitudinal analysis, we excluded self-reported T2D cases and only included 216 newly diagnosed, treatment-naïve cases (88 Chinese, 64 Malay, 64 Indian) identified at the follow-up visit and 458 controls (200 Chinese, 139 Malay, 119 Indian) who had plasma proteins measured at both time points (**Supplementary Figure 1**). The baseline and follow-up characteristics of incident T2D cases and controls, as well as the longitudinal phenotype traits changes are presented in **Supplementary Table 1**. At baseline, participants who later developed T2D had a higher BMI, waist circumference, systolic blood pressure, and TG, and lower HDL-cholesterol than the controls (all *P*<1×10^-12^). However, in the longitudinal analysis, changes in these risk factors were not associated with T2D (all *P*>0.05).

**Figure 1.**
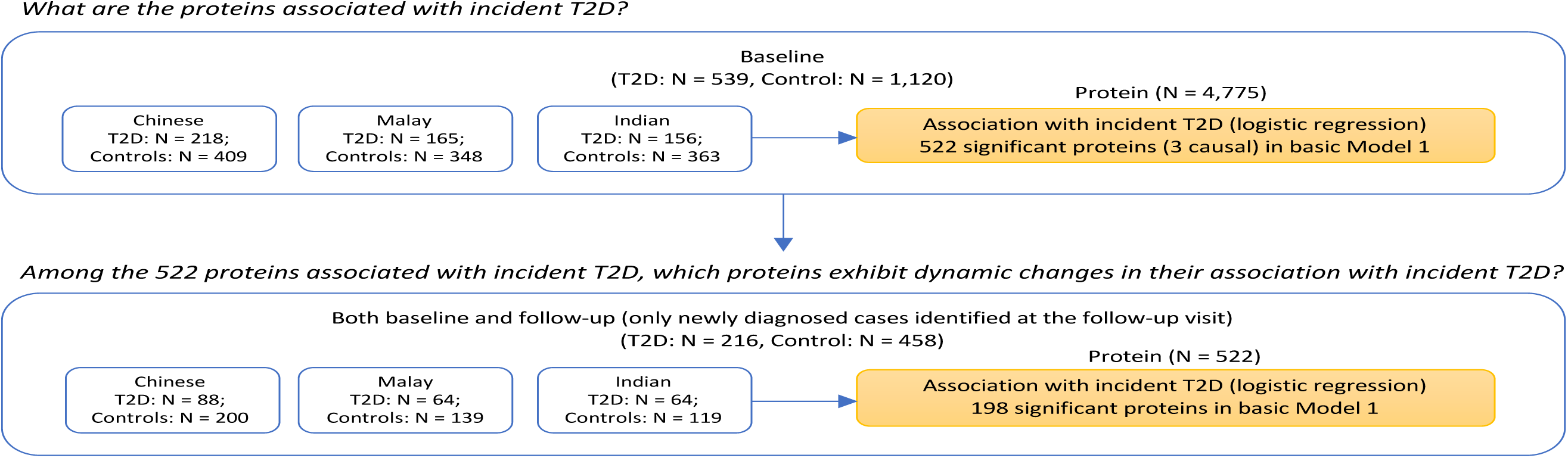
Summary of the study design, methods and primary results.

### Incident T2D association with proteins at baseline

We identified 522 proteins significantly associated with incident T2D after adjusting for age, sex, and ethnicity at baseline (Model 1; *P*_Bonferroni_<1.05×10^-5^). The strongest associations observed at *P* <1.05 × 10^-5^ included MXRA8 (OR 0.05; 95% CI 0.03 – 0.08; *P* = 6.79 × 10^-36^), ADIPOQ (OR 0.25; 95% CI 0.20 – 0.31; *P* = 1.30 × 10^-34^), SLITRK3 (OR 0.30; 95% CI 0.25 – 0.37; *P* = 1.80 × 10^-33^) and ADH4 (OR 2.12; 95% CI 1.87 – 2.41; *P* = 1.53 × 10^-29^) (Figure 2A and **Supplementary Table 2**). Among the 522 proteins identified in our study, 316 (60.64%) proteins were not previously reported to be associated with T2D risk (**Supplementary Figure 6**). We compared our findings to four published studies on proteomics and T2D incidence in European and African populations [12, 13, 15, 16]. Of the 530 proteins that were significant at a Bonferroni threshold (approximately 1×10^-5^) or had an FDR q-value less than 0.05 in at least one other study, 479 (90.38%) were present in our data. Of these, 384 (80.17% of 479) showed directionally consistent associations and 16 proteins were reported as significant in all five studies (**Supplementary Figure 6** and **Supplementary Table 3**).

**Figure 2.**
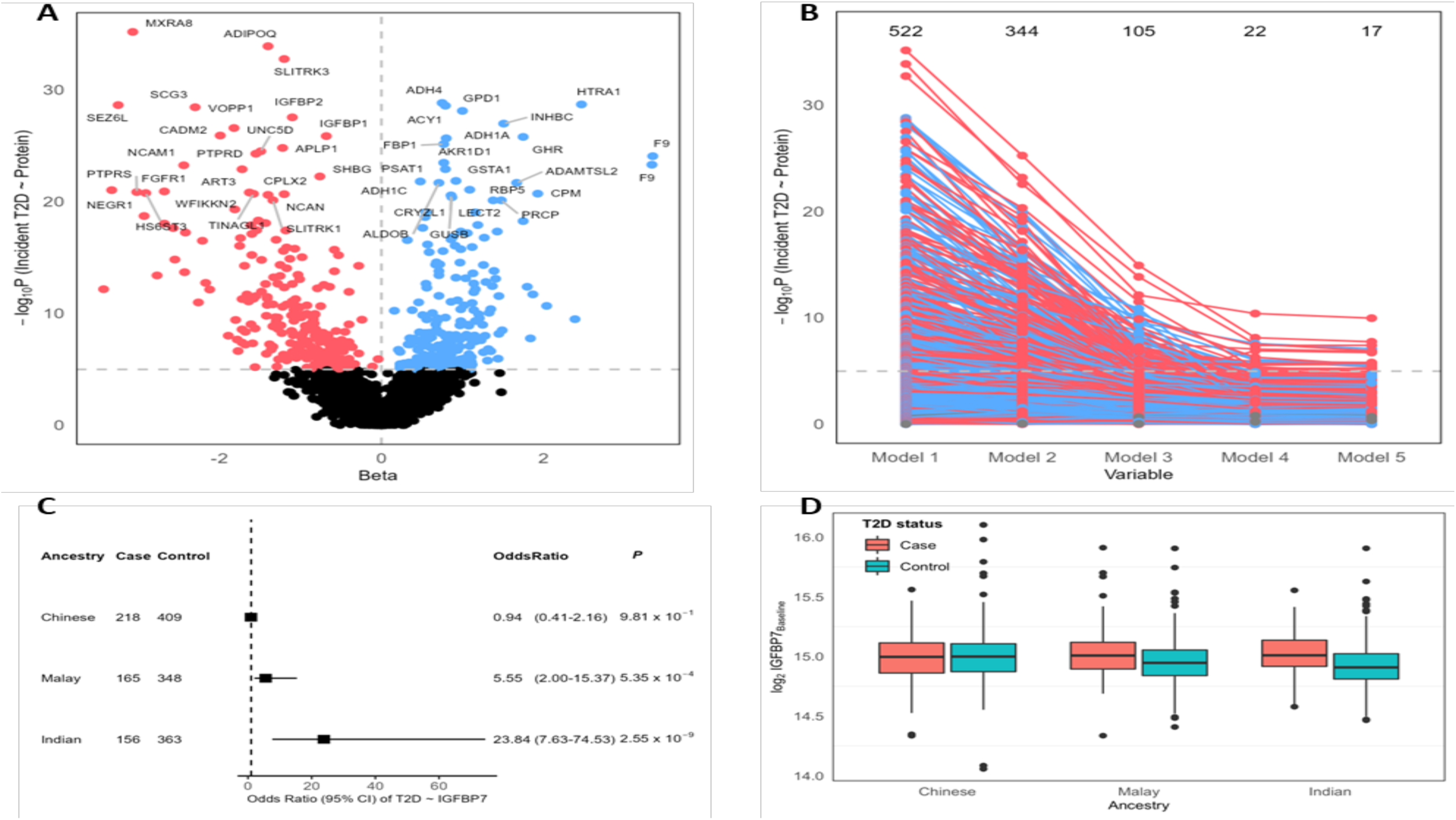
Plasma protein associations with incident type 2 diabetes (T2D). **(A)** Volcano plot of -log_10_(*P*) of plasma protein associations with incident T2D. Points representing proteins with *P*<1.05×10^-5^ are colored in blue (positive) and red (negative). Proteins with *P*<1×10^-20^ are indicated in the plot. **(B)** Spaghetti plots depicting -log_10_(*P*) trend (y axis) for protein associations with incident T2D, using five models (x axis). Model 1 includes age, sex, and ethnicity. Full Model 5 includes age, sex, ethnicity, fasting glucose (Glu), waist circumference (WC), triglycerides (TG), systolic blood pressure, and family history of diabetes. The grey dashed line represents the Bonferroni-corrected significance thresholds (*P* = 1.05 × 10^-5^). The numbers above represent the significant protein counts in each model. The color of the points/lines reflects whether the connections are positive (blue) or negative (red). **(C)** Forest plot of odds ratios, confidence intervals and *P*-values for the association between IGFBP7 and incident T2D risk in Chinese, Malay and Indian separately. **(D)** Boxplots depicting baseline IGFBP7 protein levels in T2D cases and controls across diverse ethnicity groups (Chinese, Malay, and Indian). Y-axis represents log_2_-transformed IGFBP7 protein levels at baseline. Red: T2D cases; Blue: controls.

We sequentially adjusted for additional T2D risk factors through a stepwise model selection process. After adjusting for fasting glucose, 344 proteins (65.90% of 522 in Model 1) remained significant. After further adjustment for waist circumference (105 proteins; 20.11% of 522) and both waist circumference and TG (22 proteins; 4.21% of 522), a markedly smaller number of proteins remained significant, demonstrating the substantial influence of these risk factors on the association between proteins and incident T2D (Figures 2B**; Supplementary Figure 7; Supplementary Tables 2, 4** and **5**). Specifically for waist circumference, 45.79% (N = 239) of the signals were attenuated and no longer significant when waist circumference was included in the model (**Supplementary Table 5**). Lastly, in Model 5 with additional variables SBP and family history of diabetes, 17 (3.26% of 522) proteins remained significantly associated with T2D risk at the Bonferroni-corrected threshold (**Table 1** and Figure 2B).

**Table 1.**
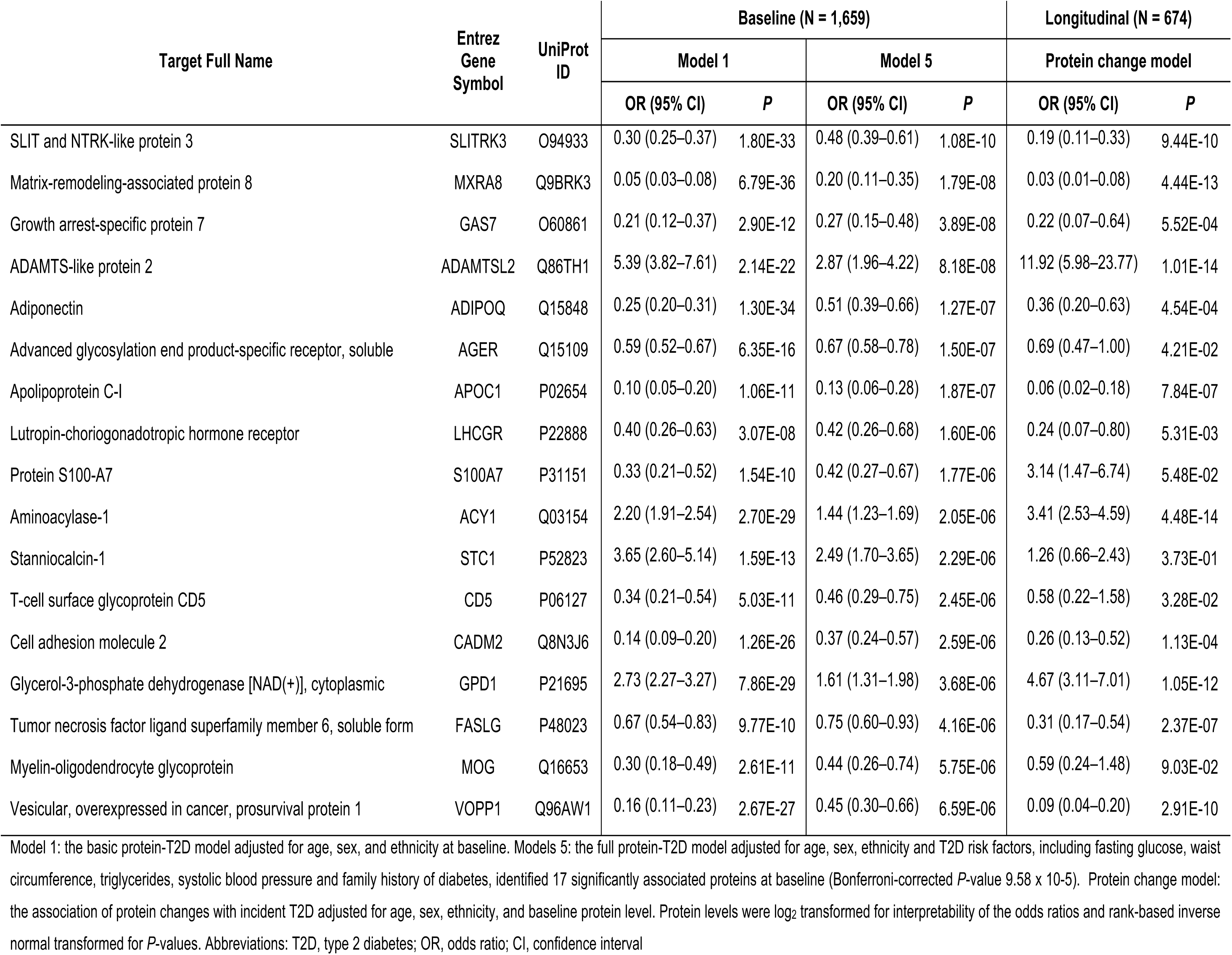
List of 17 plasma proteins associated with incident T2D in Model 5 adjusted for age, sex, ethnicity, fasting glucose (Glu), waist circumference (WC), triglycerides (TG), systolic blood pressure, and family history of diabetes at baseline in the MEC1 study.

| Target Full Name | Entrez Gene Symbol | UniProt ID | Baseline (N = 1,659) |  |  |  | Longitudinal (N = 674) |  |
| --- | --- | --- | --- | --- | --- | --- | --- | --- |
|  |  |  | Model 1 |  | Model 5 |  | Protein change model |  |
|  |  |  | OR (95% CI) | P | OR (95% CI) | P | OR (95% CI) | P |
| SLIT and NTRK-like protein 3 | SLITRK3 | O94933 | 0.30 (0.25–0.37) | 1.80E-33 | 0.48 (0.39–0.61) | 1.08E-10 | 0.19 (0.11–0.33) | 9.44E-10 |
| Matrix-remodeling-associated protein 8 | MXRA8 | Q9BRK3 | 0.05 (0.03–0.08) | 6.79E-36 | 0.20 (0.11–0.35) | 1.79E-08 | 0.03 (0.01–0.08) | 4.44E-13 |
| Growth arrest-specific protein 7 | GAS7 | O60861 | 0.21 (0.12–0.37) | 2.90E-12 | 0.27 (0.15–0.48) | 3.89E-08 | 0.22 (0.07–0.64) | 5.52E-04 |
| ADAMTS-like protein 2 | ADAMTSL2 | Q86TH1 | 5.39 (3.82–7.61) | 2.14E-22 | 2.87 (1.96–4.22) | 8.18E-08 | 11.92 (5.98–23.77) | 1.01E-14 |
| Adiponectin | ADIPOQ | Q15848 | 0.25 (0.20–0.31) | 1.30E-34 | 0.51 (0.39–0.66) | 1.27E-07 | 0.36 (0.20–0.63) | 4.54E-04 |
| Advanced glycosylation end product-specific receptor, soluble | AGER | Q15109 | 0.59 (0.52–0.67) | 6.35E-16 | 0.67 (0.58–0.78) | 1.50E-07 | 0.69 (0.47–1.00) | 4.21E-02 |
| Apolipoprotein C-I | APOC1 | P02654 | 0.10 (0.05–0.20) | 1.06E-11 | 0.13 (0.06–0.28) | 1.87E-07 | 0.06 (0.02–0.18) | 7.84E-07 |
| Lutropin-choriogonadotropic hormone receptor | LHCGR | P22888 | 0.40 (0.26–0.63) | 3.07E-08 | 0.42 (0.26–0.68) | 1.60E-06 | 0.24 (0.07–0.80) | 5.31E-03 |
| Protein S100-A7 | S100A7 | P31151 | 0.33 (0.21–0.52) | 1.54E-10 | 0.42 (0.27–0.67) | 1.77E-06 | 3.14 (1.47–6.74) | 5.48E-02 |
| Aminoacylase-1 | ACY1 | Q03154 | 2.20 (1.91–2.54) | 2.70E-29 | 1.44 (1.23–1.69) | 2.05E-06 | 3.41 (2.53–4.59) | 4.48E-14 |
| Stanniocalcin-1 | STC1 | P52823 | 3.65 (2.60–5.14) | 1.59E-13 | 2.49 (1.70–3.65) | 2.29E-06 | 1.26 (0.66–2.43) | 3.73E-01 |
| T-cell surface glycoprotein CD5 | CD5 | P06127 | 0.34 (0.21–0.54) | 5.03E-11 | 0.46 (0.29–0.75) | 2.45E-06 | 0.58 (0.22–1.58) | 3.28E-02 |
| Cell adhesion molecule 2 | CADM2 | Q8N3J6 | 0.14 (0.09–0.20) | 1.26E-26 | 0.37 (0.24–0.57) | 2.59E-06 | 0.26 (0.13–0.52) | 1.13E-04 |
| Glycerol-3-phosphate dehydrogenase [NAD(+)], cytoplasmic | GPD1 | P21695 | 2.73 (2.27–3.27) | 7.86E-29 | 1.61 (1.31–1.98) | 3.68E-06 | 4.67 (3.11–7.01) | 1.05E-12 |
| Tumor necrosis factor ligand superfamily member 6, soluble form | FASLG | P48023 | 0.67 (0.54–0.83) | 9.77E-10 | 0.75 (0.60–0.93) | 4.16E-06 | 0.31 (0.17–0.54) | 2.37E-07 |
| Myelin-oligodendrocyte glycoprotein | MOG | Q16653 | 0.30 (0.18–0.49) | 2.61E-11 | 0.44 (0.26–0.74) | 5.75E-06 | 0.59 (0.24–1.48) | 9.03E-02 |
| Vesicular, overexpressed in cancer, prosurvival protein 1 | VOPP1 | Q96AW1 | 0.16 (0.11–0.23) | 2.67E-27 | 0.45 (0.30–0.66) | 6.59E-06 | 0.09 (0.04–0.20) | 2.91E-10 |

Model 1: the basic protein-T2D model adjusted for age, sex, and ethnicity at baseline. Models 5: the full protein-T2D model adjusted for age, sex, ethnicity and T2D risk factors, including fasting glucose, waist circumference, triglycerides, systolic blood pressure and family history of diabetes, identified 17 significantly associated proteins at baseline (Bonferroni-corrected $P$ -value $9.58 \times 10^{-5}$ ). Protein change model: the association of protein changes with incident T2D adjusted for age, sex, ethnicity, and baseline protein level. Protein levels were $\log_2$ transformed for interpretability of the odds ratios and rank-based inverse normal transformed for $P$ -values. Abbreviations: T2D, type 2 diabetes; OR, odds ratio; CI, confidence interval

To assess possible differential effects on T2D across different Asian ethnicity groups, we examined the interaction between 4,775 proteins and ethnicity (Chinese, Malay, and Indian) in association with incident T2D. We found a significant interaction between the IGFBP7 and ethnicity (*P*_interaction_ = 7.35 × 10^-6^) in the model adjusted for age, sex, and ethnicity (**Supplementary Table 2**). Further analyses stratified by ethnicity suggested that the interaction was mainly driven by the Indian participants (Indian: OR 23.84, 95% CI 7.63 – 74.53; *P* = 2.55 × 10^-9^; Malay: OR 5.55; 95% CI 2.00 – 15.37; *P* = 5.35 × 10^-^

^4^; Chinese: OR 0.94, 95% CI 0.41 – 2.16; *P* = 9.81 × 10^-1^) (Figures 2C and **2D**). We note the wide confidence interval as a result of smaller sample sizes for the strata. The lead variant (rs1543178, *P* = 1.48 × 10^-6^) associated with IGFBP7 in the pQTL meta-analysis exhibited variation in minor allele frequency across these ethnic groups: 0.01 in Chinese, 0.05 in Malay, and 0.12 in Indian (**Supplementary Figure 8**)

### Change in plasma proteins and T2D

We further assessed the longitudinal change of the 522 incident T2D-associated protein biomarkers in relation to T2D onset. In 216 treatment-naïve cases based on glycemic measures at follow-up and 458 controls, changes in the levels of 205 (39.27%) proteins were significantly associated with T2D adjusted for age, sex, ethnicity, and baseline protein level (*P* < 9.58 × 10^-5^ = 0.05/522) (**Supplementary Figure 9A**). Consistent direction of association was observed in 96.10% (197 out of 205) of these proteins with Pearson correlation coefficient of 0.76 (**Supplementary Figure 9B**). Adjusting for baseline fasting glucose in the longitudinal analysis reduced the number of significant associations from 205 to 180 (**Supplementary Table 5**). Among these 180 proteins, 125 (69.44%) were associated with differences in fasting glucose measured at two time points (**Supplementary Table 4**). Finally, 178 proteins remained associated with in the full model with additional adjustment for baseline fasting glucose, family history of diabetes at baseline, and changes in waist circumference, TG, and SBP (**Supplementary Tables 5** and **6**).

### Functional enrichment analysis

The 522 proteins associated with incident T2D were enriched for pathways, including insulin-like growth factor binding, glycosaminoglycan binding, and hormone metabolic processes (**Supplementary Figure 10A** and **Supplementary Table 7**). While the functional enrichment analysis suggested enrichment in the liver, consistent with previous research in western populations (**Supplementary Figure 10B**) [12], we note that proteins included on the SomaScan panel may be enriched in the liver (**Supplementary Note**). Similar enriched pathways were also observed for the subset of 205 proteins that changed significantly between baseline and follow-up (**Supplementary Figures 10C, 10D** and **11**).

### Evaluating causal relationships between proteins and T2D

Using two-sample MR analysis to evaluate the causal relationship between proteins and T2D, only 26.63% (N = 139) of the 522 incident T2D-associated proteins had valid genetic instruments for proteins and summary statistics available from Mahajan et al. [9] (**Supplementary Note**). In the two-sample MR analysis, three proteins (GSTA1, INHBC, and FGL1) had evidence (*P* _FDR_ < 0.05) of a causal effect on T2D (**Table 2** and **Supplementary Table 8**). Directions of effect were consistent between causal and observational estimates and both incident and longitudinal analysis (**Table 2**). GSTA1 has previously been reported to be causally associated with T2D, while INHBC was identified as a consequence of T2D [12, 13]. Colocalization analysis between pQTL instruments and external T2D GWAS showed evidence of colocalization for GSTA1 (PP H4 = 83.70%) and INHBC (PP H4 = 71.20%) but not FGL1 (PP H4 = 1.10%) (**Supplementary Table 8** and **Supplementary Figure 12**).

**Table 2.** Proteins with evidence of causal associations with T2D in MR analysis.

| Protein Name | Protein | MR analysis |  |  |  |  |  | Colocalization analysis | Association with Incident T2D * |  |  |  |
| --- | --- | --- | --- | --- | --- | --- | --- | --- | --- | --- | --- | --- |
|  |  |  |  |  |  |  |  |  | Baseline protein (Model 1) |  | Protein change |  |
|  |  | Method | IV (N) | Beta | SE | P | FDR.P | PP H4 | Beta | P | Beta | P |
| Glutathione S-transferase A1 | GSTA1 | Wald ratio | 1 | 0.05 | 0.01 | 4.04 x 10<sup>-5</sup> | 6.95 x 10<sup>-3</sup> | 83.70% | 0.92 | 1.44 x 10<sup>-22</sup> | 1.08 | 6.00 x 10<sup>-8</sup> |
| Inhibin beta C chain | INHBC | Wald ratio | 1 | 0.13 | 0.03 | 2.97 x 10<sup>-4</sup> | 2.55 x 10<sup>-2</sup> | 71.20% | 1.51 | 1.08 x 10<sup>-27</sup> | 2.6 | 1.50 x 10<sup>-10</sup> |
| Fibrinogen-like protein 1 | FGL1 | Wald ratio | 1 | -0.06 | 0.02 | 6.17 x 10<sup>-4</sup> | 3.54 x 10<sup>-2</sup> | 1.10% | -0.39 | 2.35 x 10<sup>-6</sup> | -0.39 | 4.50 x 10<sup>-2</sup> |

\*Both incident T2D association models were adjusted for age, sex and ethnicity. Abbreviations: T2D, type 2 diabetes; MR, Mendelian randomization; IV (N), number of instrument variants; SE, standard error; FDR.P, false discovery rate corrected *P*-value; PP H4: posterior probability of hypothesis 4. Proteins with significance levels of FDR corrected *P* < 0.05 are displayed here, while the remaining proteins are listed in Supplementary Table 7.

## Discussion

We measured 4,775 proteins in 1,659 participants (539 incident cases and 1120 controls) of Chinese, Indian, and Malay ethnicity with the SomaScan assay. Over 79% of previously reported associated proteins (n = 479) showed a consistent direction of effect in our study, representing a validated set of diabetes-associated proteins that were replicated across geographically and diverse populations [12, 13, 15, 16]. A test of heterogeneity identified a single protein (IGFBP7) for which the association with T2D was modified by ethnicity. However, we recognize that our study had limited power to detect interactions. These findings suggest that many of the biological processes in the pathophysiology of T2D (those captured by measurements of plasma proteins) are shared across ancestry groups despite apparently heterogeneous T2D phenotype manifestations.

We identified 522 proteins associated with incident T2D at the Bonferroni-corrected threshold, including 316 that have not been previously reported to be associated with T2D. Many of these novel associations may have arisen due to the larger number of proteins assayed in our study than earlier versions of the SomaScan platform. We also reported associated proteins with basic age, sex and ethnicity adjustment In some previous studies, models adjusted for other physiologic factors known to affect plasma protein levels that may be altered early in the natural history of disease, particularly in T2D with a prolonged natural history were reported [12, 13, 15, 31]. It is commonly accepted that obesity and insulin resistance are early events in the pathogenesis of T2D, with beta-cell dysfunction leading to hyperglycemia at a later stage [3]. Our data showed that (i) excess adiposity and elevated triglyceride (which are markers for insulin resistance) were already observed at baseline, and (ii) adjustment for adiposity and triglycerides attenuated protein-T2D associations reflecting correlations of proteins with these risk factors. The three proteins identified by MR analysis were also associated with baseline fasting glucose and TG, and their association significance was attenuated after adjustment for these factors. This highlights the limitations of observational analysis to distinguish between confounding and mediation by other biological risk factors and the value of MR and colocalization analyses to provide evidence on causality.

The proteins associated with incident T2D and those exhibiting associations across baseline and follow-up longitudinal change with incident T2D were involved in metabolism and the insulin response system, and enriched for liver-specific gene expression, which was consistent with previous studies in western populations using the SomaScan panel [12, 13]. After adjustment for other known risk factors for T2D, 17 proteins remained associated with incident T2D, of which seven proteins (STC1, GAS7, APOC1, MOG, CD5, S100A7, and LHCGR) were novel. While these protein biomarkers will likely provide limited predictive gain over more conventional biomarkers such as obesity and fasting plasma glucose, omics biomarkers associated with T2D have the potential to provide insight into the biological pathways involved in the pathophysiology linking known risk factors with T2D development and potentially to the complications of T2D. For example, STC1 receptors are found in mouse pancreatic beta calls and that its ligand co-localizes with insulin [32], and APOC1 helps prevent insulin resistance by reducing fat storage, even when plasma triglyceride levels are high in experimental studies in mice [33]. Insulin-like growth factor-binding protein 7 (IGFBP7) is the only protein that showed an interaction with ethnicity in its association with T2D. IGFBP7, known for its growth-suppressing properties, plays a key role in protein synthesis, cell growth, and cell survival [34]. This protein’s strong binding affinity for insulin suggests its potential role in insulin resistance in T2D [35–37]. Further research is needed to evaluate whether these novel protein biomarkers for T2D risk may result from genetic or lifestyle differences between different populations.

Mendelian randomization identified three proteins (GSTA1, INHBC, and FGL1) that may have a casual role in T2D development. Both GSTA1 and INHBC exhibited consistent directions of effect in both causal and observational estimates, further substantiated by colocalization evidence within our dataset. Limited by a small sample size, none of the three proteins had more than two independent genetic instruments that would allow us to test for pleiotropy. We replicated the finding of the causal role of GSTA1 in T2D from the U.S. ARIC study. GSTA1 (Glutathione S-transferase A1), prominently expressed in liver and kidney tissues, functions as an indicator of hepatic function and plays a pivotal role in the body’s antioxidant system by facilitating the binding of glutathione to carcinogens, drugs, and oxidative stress byproducts in mice [38]. Inhibin Beta C Chain (INHBC, a protein of the TGF-β family) is secreted by the liver and regulates the secretion of gonadal hormones and insulin [39, 40]. However, MR analysis in the AGES-Reykjavik study suggested that altered concentrations of INHBC were a consequence and not a cause of T2D, and that it is associated with both incident and prevalent T2D. We note that INHBC showed significant changes between baseline and follow-up in our data. No causal effects on T2D have previously been reported for FGL1, but it was found to promote adipogenesis in animal and cell models [41]. None of these three proteins remain significant after adjustment for other risk factors. These protein associations with T2D risk were primarily accounted for by specific adjustments: FGL1 by fasting glucose adjustment, GSTA1, and INHBC by triglyceride adjustment. These findings imply that glucose metabolism, lipid metabolism, and abdominal obesity may mediate the causal link between these proteins and T2D. To bolster the robustness and reliability of our findings, additional validation with larger sample sizes is needed.

Our study has several limitations. First, we did not validate our findings in independent Asian populations due to the lack of other Asian studies with data on proteomics and T2D incidence. Prioritizing larger Asian population sets in future proteomics research can improve the robustness and transferability of findings. Second, the prolonged natural history of T2D can affect protein levels even in prospective studies, as highlighted by the poorer metabolic risk profile prior to T2D diagnosis. Third, the presence of different significance levels among different aptamers targeting the same protein in our analysis suggests differences in the capture specificity of the assay with a median correlation of about 0.4 observed for overlapping proteins between SomaLogic and Olink platforms as previously reported [42–44]. Therefore, future research should explore combining results across panels. Finally, while we observed an enrichment for liver-specific transcripts from the 522 incident-T2D associated proteins, other organ systems involved in T2D such as the beta-cell may not secrete significant amounts of protein to be detected using plasma proteomics. This underscores a potential limitation on plasma proteomics panels for understanding disease pathophysiology, emphasizing the need for caution in interpretation and consideration of complementary approaches.

In conclusion, we have identified multiple plasma proteins, including 316 novel associations, associated with incident T2D in a population of diverse Asian ancestry. Our data suggests that many of the pathophysiologic pathways leading to T2D are likely shared between populations of different ancestry. Plasma proteomics is particularly well suited to detect changes in proteins secreted by the liver and for T2D, particularly those associated with obesity, insulin resistance and glucose metabolism disorder (even relatively mild changes that occur well before the onset of T2D). Our findings on these plasma proteins provide evidence for causality and implicate novel biological pathways associated with the identified protein risk factors, which could be targeted by future preventive interventions.

## Data Availability

Summary statistics for all measured proteins are provided in the Supplementary Tables. Data from the Singapore Multi-Ethnic Cohort Phase 1 study can be requested by researchers for scientific purposes through an application process at the listed website (https://blog.nus.edu.sg/sphs/data-and-samples-request/). Data will be shared through an institutional data sharing agreement.

## Author Contributions

X.S., R.M.vD. and E.S.T. designed the study. Y.L., C.G.Y.L., S.C.R., N.B., J.C, J.Y., Yun.L. and E.S.T., R.M.vD., X.S. contributed to the analysis and interpretation of data. Y.L., X.S., E.S.T. and R.M.vD. wrote the draft of the manuscript. All authors contributed to critical revision of the manuscript.

## Disclaimers

The National University of Singapore has signed a collaboration agreement with SomaLogic to conduct SomaScan of Singapore Multi-Ethnic Cohort Phase 1 (MEC1) stored samples at no charge in exchange for the rights to analyze linked MEC1 phenotype data.

## Acknowledgements

We thank all participants, the study team, and the investigators for their research contributions. The MEC1_T1 and MEC1_T2 studies are supported by individual research and clinical scientist award schemes from the National Medical Research Council (NMRC) and the Biomedical Research Council (BMRC) of Singapore, and infrastructure funding from the Singapore Ministry of Health (Population Health Metric and Analytics PHMA), National University of Singapore and National University Health System, Singapore. This study made use of whole-genome data generated on MEC1 as part of the Singapore National Precision Medicine program funded by the Industry Alignment Fund (Pre-Positioning) (IAF-PP: H17/01/a0/007)

S.C.R was supported by core funding from the: British Heart Foundation (RG/18/13/33946), the Munz Chair of Cardiovascular Prediction and Prevention and the NIHR Cambridge Biomedical Research Centre (NIHR203312) [*], Cambridge BHF Centre of Research Excellence (RE/18/1/34212) and BHF Chair Award (CH/12/2/29428) and by Health Data Research UK, which is funded by the UK Medical Research Council, Engineering and Physical Sciences Research Council, Economic and Social Research Council, Department of Health and Social Care (England), Chief Scientist Office of the Scottish Government Health and Social Care Directorates, Health and Social Care Research and Development Division (Welsh Government), Public Health Agency (Northern Ireland), British Heart Foundation and Wellcome. *The views expressed are those of the authors and not necessarily those of the NIHR or the Department of Health and Social Care.

## Supplementary Figures

**Supplementary Figure 1.**
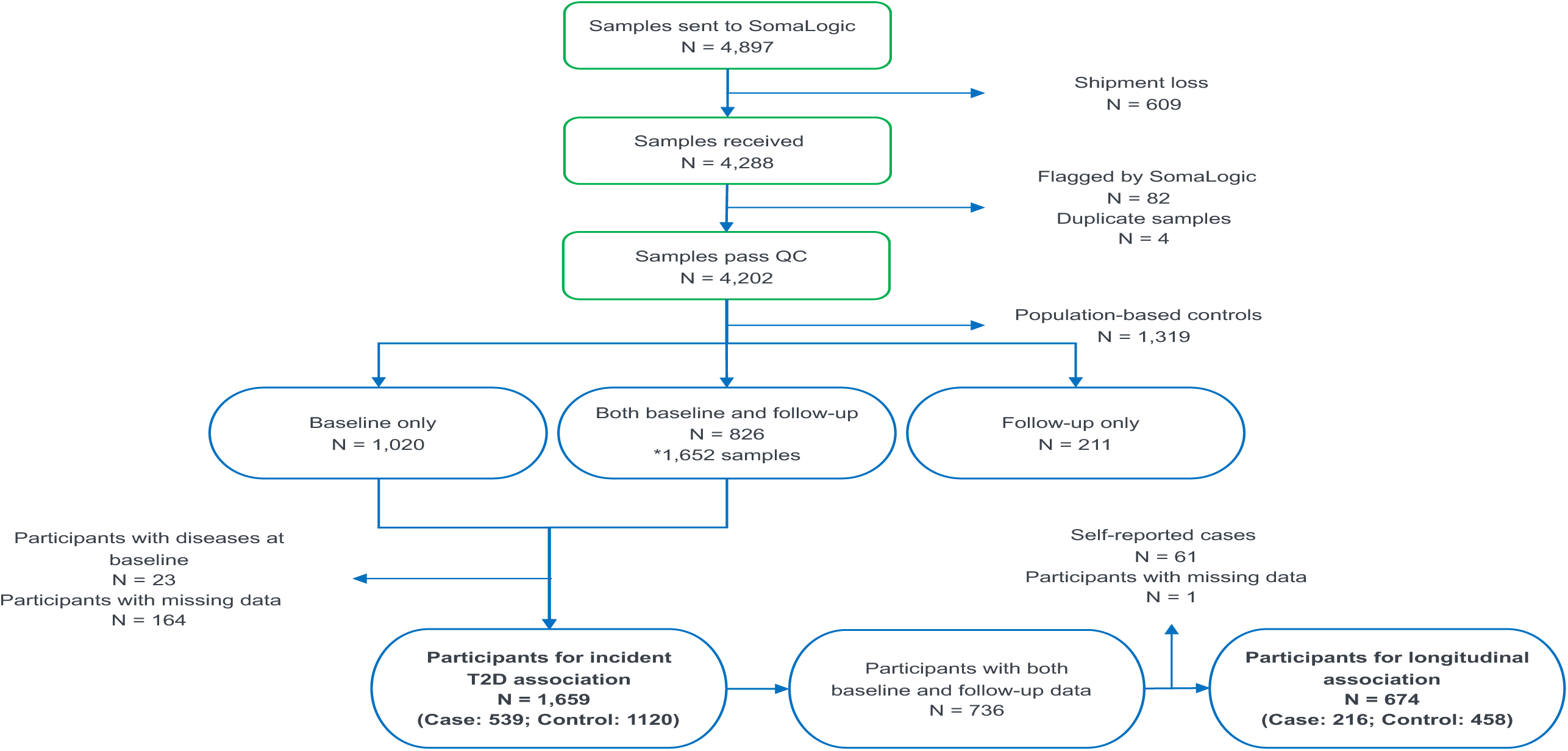
Flow chart illustrating the inclusion and exclusion of samples (at a single timepoint) or participants (with single or two timepoints) into the study. Green rectangles denote sample counts and blue ovals denote participant counts.

**Supplementary Figure 2.**
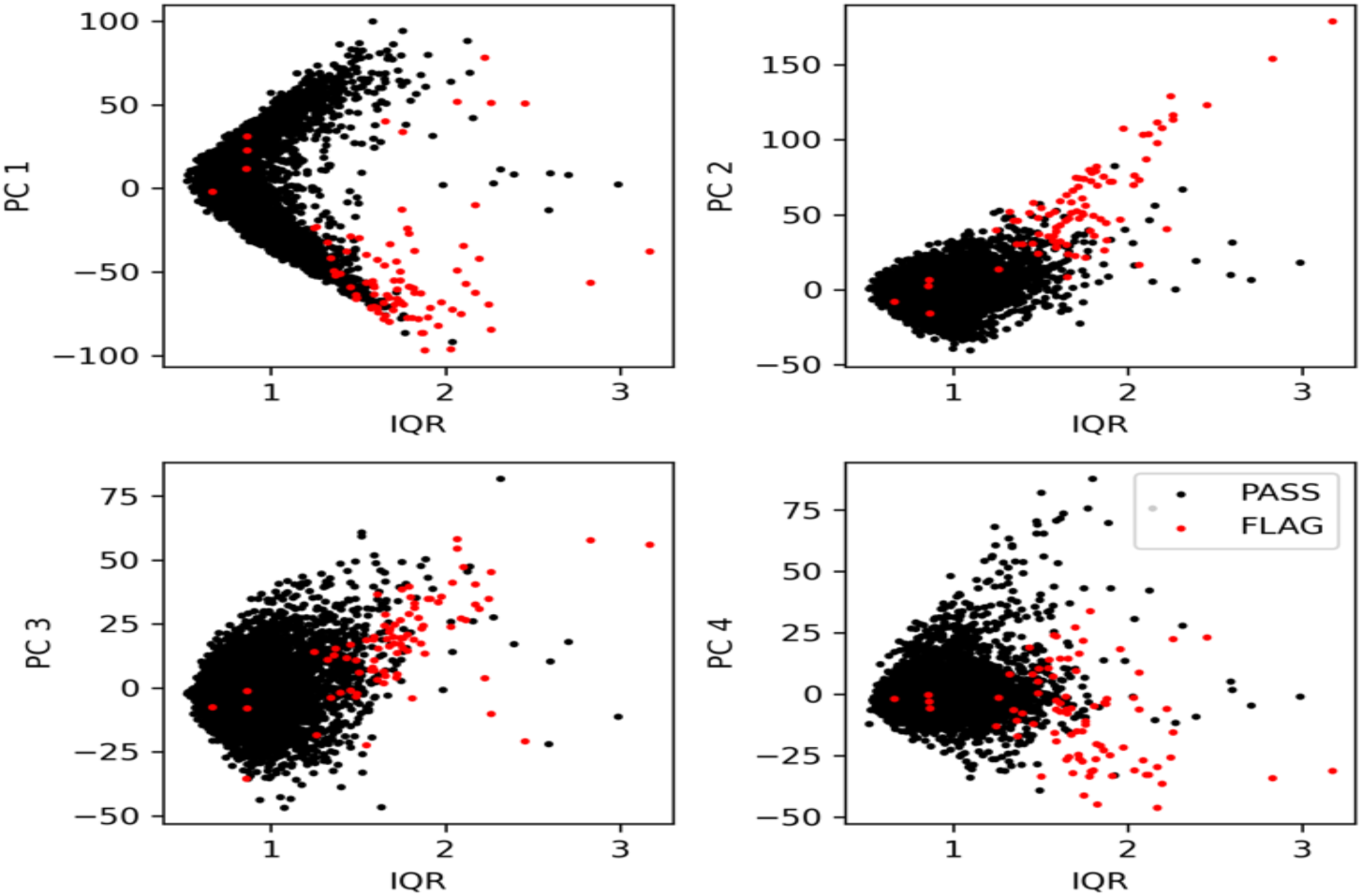
Principal components (PC) from principal component analysis plotted against interquartile range (IQR) of 4,288 samples based on log_2_-transformed aptamer levels. 82 samples flagged by SomaLogic and excluded in our analysis are colored in red. 4,206 samples which passed SomaLogic QC are colored in black.

**Supplementary Figure 3.**
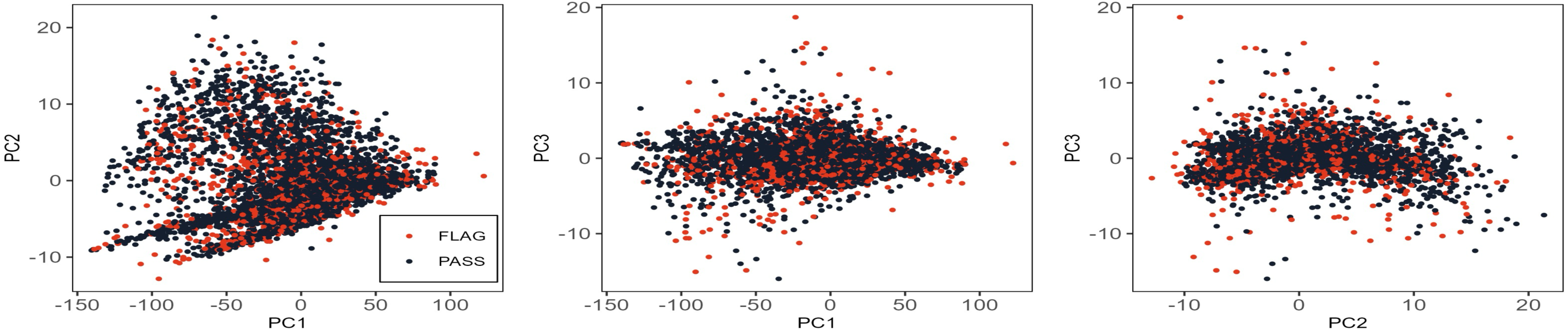
Principal component (PC) plots from principal analysis of 4,978 aptamers. Additional quality control of PCA based on log_2_-transformed aptamer levels did not exclude any aptamers. Aptamers flagged by SomaLogic but included in our analysis were colored in red. Aptamers which passed SomaLogic QC are colored in black.

**Supplementary Figure 4.**
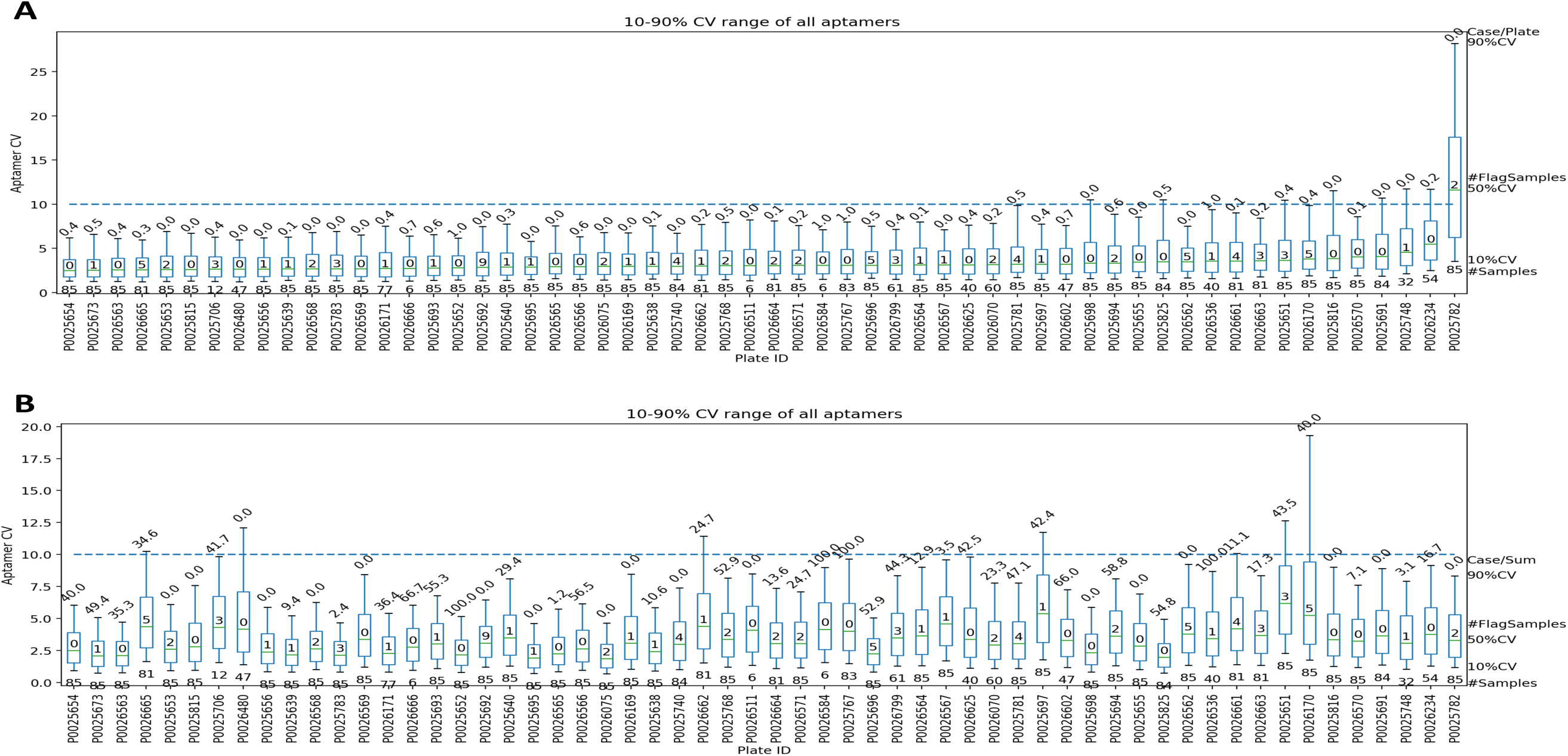
Aptamer coefficient of variations (CVs) in **(A)** Calibrator Control and **(B)** Quality Control samples across 53 plates. The median line in each box represents the 50^th^ percentile of the CV values. The upper whisker and the lower whisker represent the 90^th^ and 10^th^ percentile of the CV values, respectively. The number below the lower whisker is the total sample number in each plate and the ratio above the upper whisker is the percentage of T2D incident case plated in each plate. The dashed line is the acceptance criteria (10%) for aptamer CV.

**Supplementary Figure 5.**
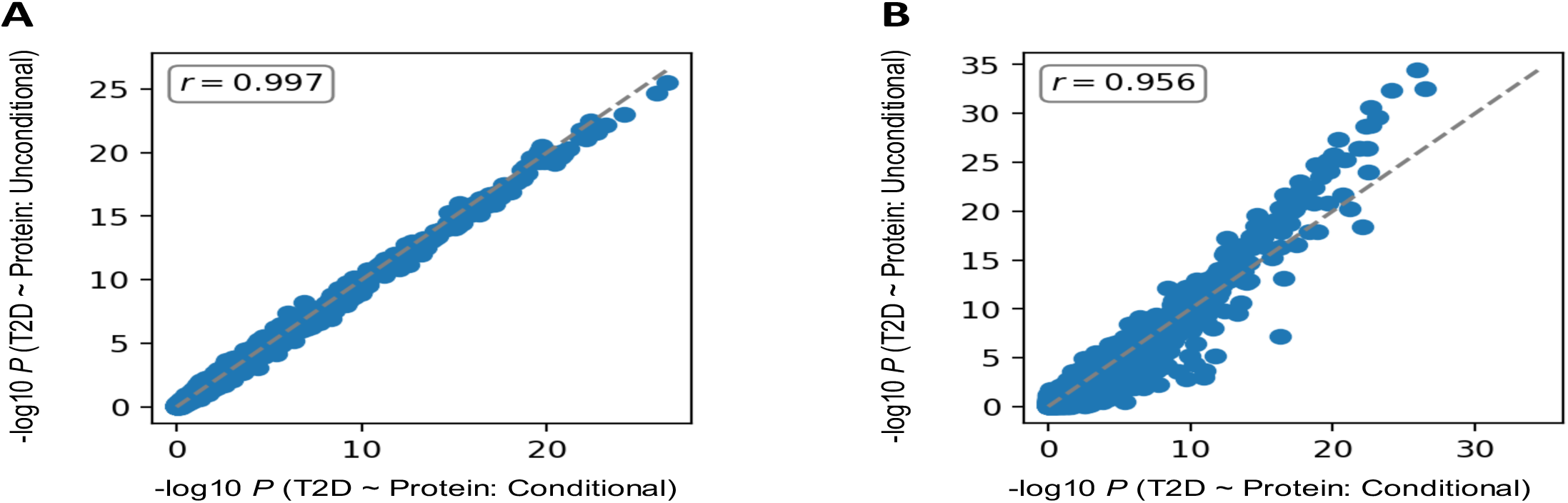
Comparison of -log_10_(*P*) for incident type 2 diabetes (T2D) association with proteins between unconditional (ignoring case/control pairing) and unconditional (accounting for case/control pairing) logistic models. **(A)** Using the same set of samples profiled under the original case/control pairing (513 cases and 789 controls), compare unconditional and conditional logistic regression models. Pearson correlation coefficient = 0.997 **(B)** Using the same conditional case/control pairing (513 cases and 789 controls), compare with unconditional logistic regression model which maximized all available sample (539 cases and 1,120 controls). Pearson correlation coefficient = 0.956. The blue points represent the 4,775 proteins. Both models were adjusted for age, sex and ethnicity.

**Supplementary Figure 6.**
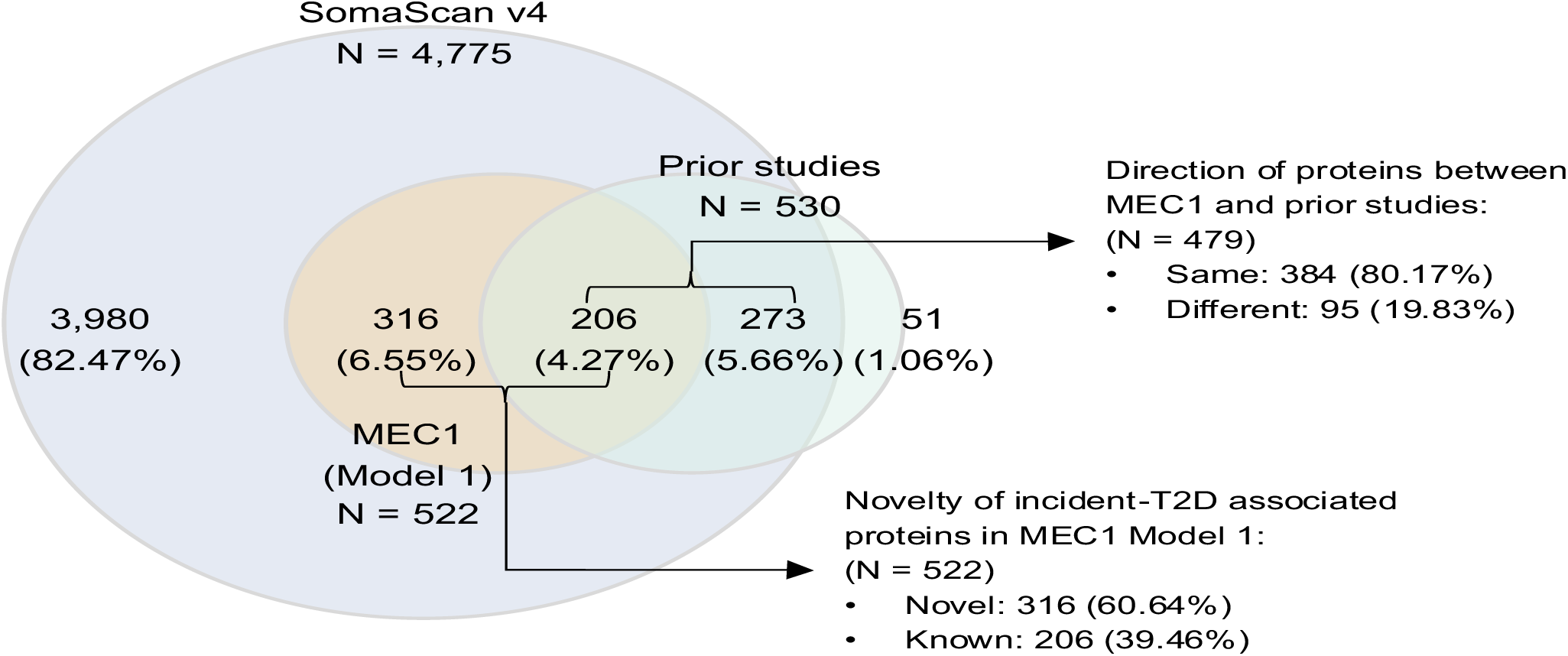
Venn diagram detailing the overlap of proteins associated with incident T2D in MEC1 and prior studies. The blue circle represents unique proteins of the SomaScan v4 panel. The yellow circle denotes proteins significantly associated with incident T2D in MEC1 Model 1 adjusting for age, sex and ethnicity, while the green circle denotes proteins identified in prior studies, including AGES-Reykjavik Study (PMID: 32385057), Atherosclerosis Risk in Communities (ARIC; PMID: 36706097), Jackson Heart Study (PMID: 36630488), Framingham Heart Study (FHS; PMID: 33591955), and Malmö Diet and Cancer Study (MDCS; PMID: 33591955). Proteins (N = 51) that do not exist in the MEC1 proteomic data were from older versions of the SomaScan panel used in previous studies. Associations between protein levels and incident T2D were identified through logistic regression in all studies with comparable basic models: MEC1 (Model 1) adjusted for age, sex, and ethnicity; AGES-Reykjavik adjusted for age and sex; ARIC adjusted for age, sex, race, and eGFR; Jackson Heart Study adjusted for age, sex, and sample batch; FHS and MDCS adjusted for age, sex, and sample batch. Percentages in the circles are calculated based on the union set of all proteins (N = 4,826).

**Supplementary Figure 7.**
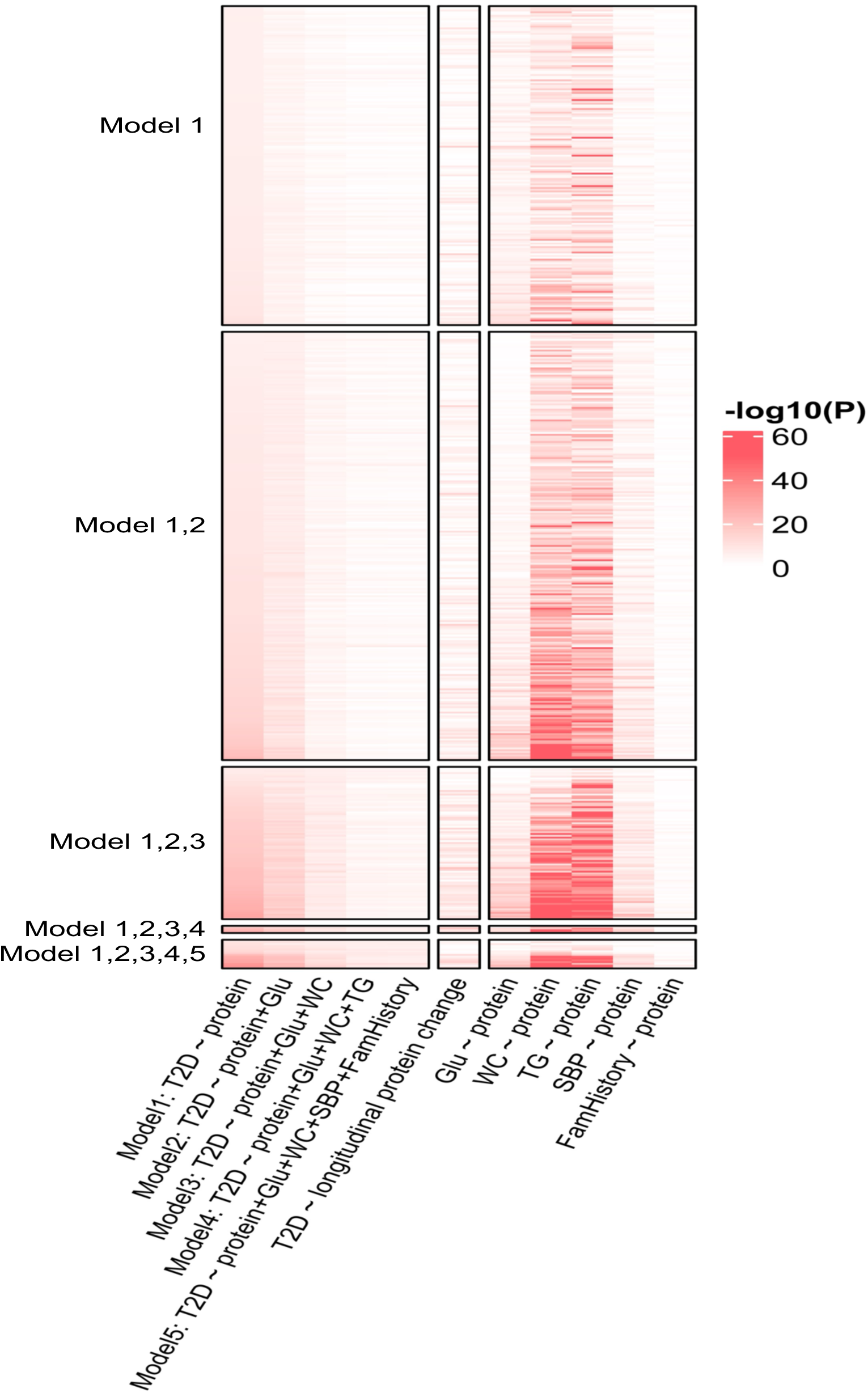
Heatmap showing significance of associations for 522 incident T2D-associated proteins with T2D and different phenotypic traits at baseline. The intensity scale in red on the heatmap represents -log_10_(*P*) association. The x-axis indicates proteins association with T2D adjusted with different phenotypic traits (Glu, WC, TG, SBP, and family history of diabetes) and proteins association with these phenotypic traits. The leftmost five columns represent Models 1 to 5 of protein association with incident T2D. Model 1, protein-T2D model adjusted for age, sex, and ethnicity that identified 522 significantly associated proteins (Bonferroni-corrected *P* = 1.05 × 10^-5^). Models 2 to 5 sequentially adjust for additional T2D risk factors, including Glu, WC, TG, SBP, and family history of diabetes. The middle column represents the model analyzing longitudinal protein changes associated with incident T2D, adjusted for age, sex, ethnicity, and baseline protein level. The other five models analyze protein associations with additional T2D risk factors: Glu, WC, TG, SBP, and family history of diabetes. The 522 associated proteins from Model 1 were clustered according to their significance across Models 2 to 5. For example, ‘Model 1’ includes proteins significant only in Model 1. ‘Model 1, 2, 3, 4, 5’ indicates proteins that remained significant in all five models after adjusting for the five key T2D risk factors. Within each subgroup, proteins are ordered by their *P*-values according to the association analysis from Model 1. T2D: type 2 diabetes; Glu: fasting glucose; WC: waist circumference; TG: triglycerides; SBP: systolic blood pressure; FamHistory: family history of diabetes.

**Supplementary Figure 8.**
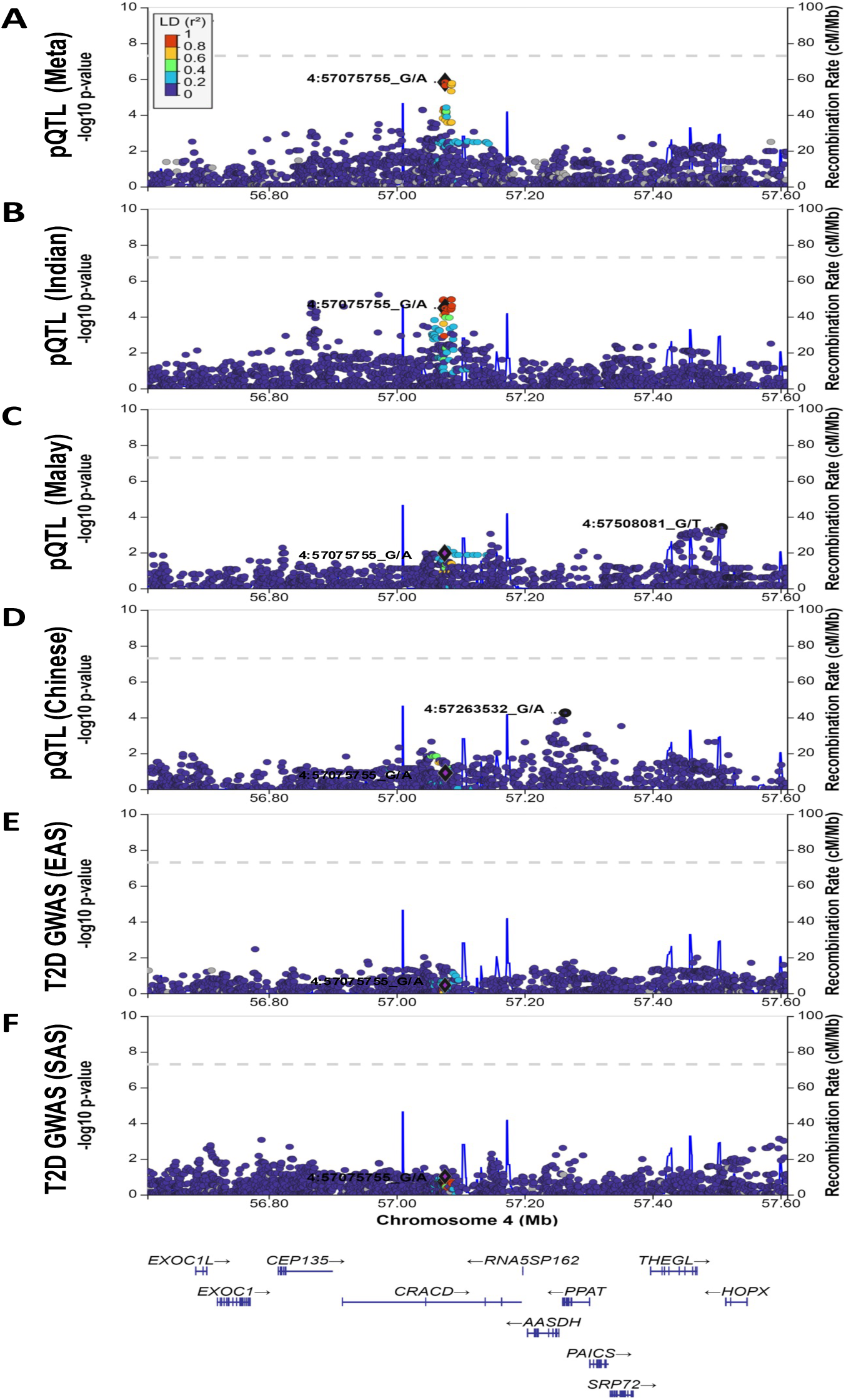
Regional association plots with IGFBP7 protein levels or type 2 diabetes (T2D) spanning ±500 kb around lead genetic variant rs1543178. Genetic associations with IGFBP7 protein in the **(A)** multi-ancestry meta-analysis across Indian, Malay, and Chinese populations, and separately in **(B)** Indian, **(C)** Malay, and **(D)** Chinese populations. Genetic associations with T2D (PMID: 35551307) in **(E)** East Asian and **(F)** South Asian populations. The lead variant rs1543178 (4:57075755_G/A) associated with IGFBP7 in the multi-ancestry meta-analysis is highlighted in purple. Lead variants from the Malay and Chinese populations are labelled within 500 Mb region of rs1543178. Grey dashed line represents genome-wide significance threshold (*P* = 5 × 10^−8^). Variants are colored based on MEC1 Indian population linkage disequilibrium with rs1543178. Genes in the region are displayed below the plots. pQTL: protein quantitative trait locus; Mb:megabase; cM:centimorgan; EAS, East Asian; SAS, South Asian.

**Supplementary Figure 9.**
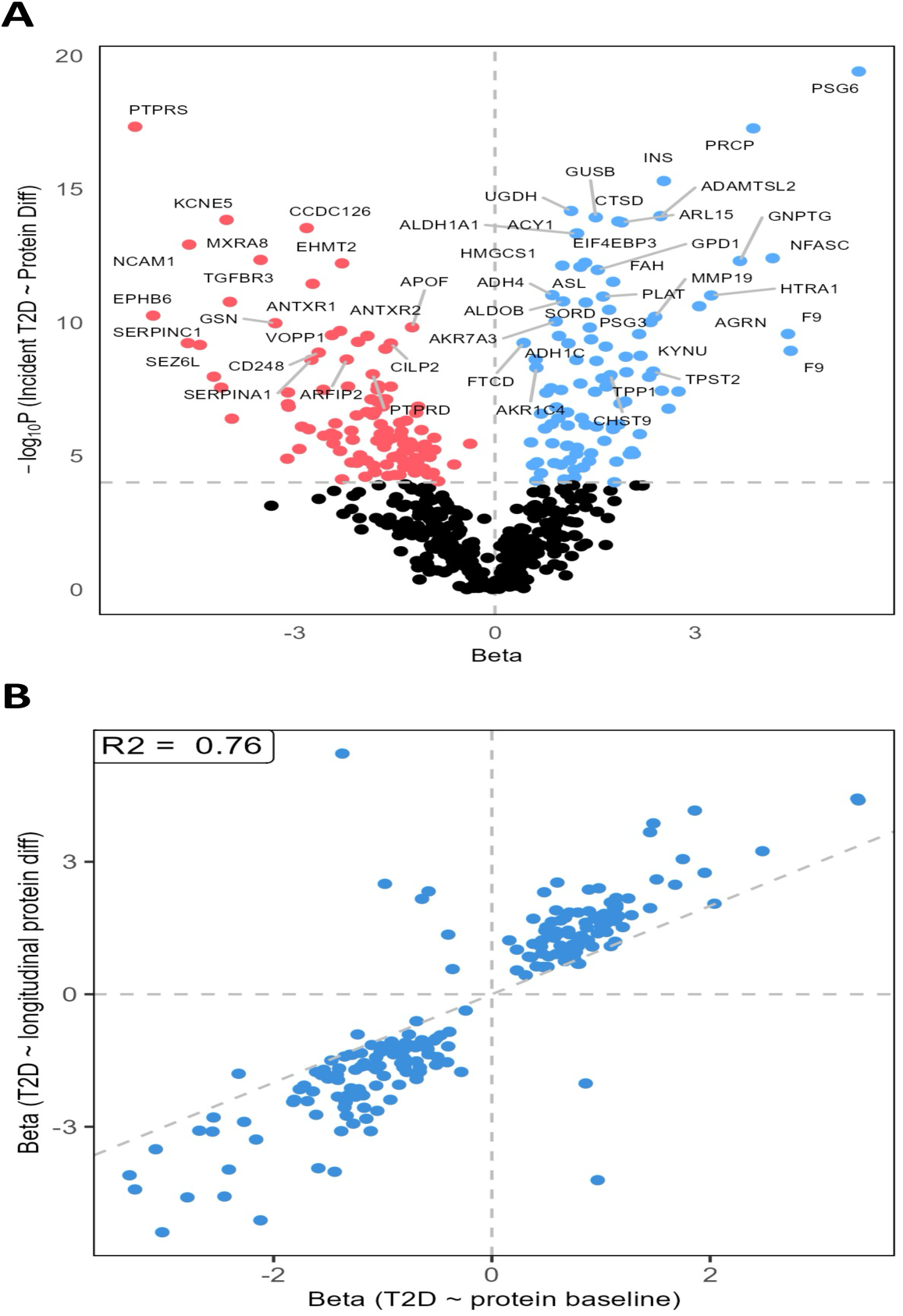
Associations of longitudinal changes in proteins with onset of type 2 diabetes (T2D) for the 522 proteins which showed an association with incident T2D. **(A)** Volcano plot of -log_10_(*P*) associations between protein changes and T2D onset. The x-axis represents the effect size generated from the association with log_2_ transformed protein levels, while the y-axis represents the -log10(*P*) generated from the association with rank-based inverse normal transformed protein levels. Proteins with *P* < 1.05 × 10^-5^ are colored blue for positive associations and red for negative associations. **(B)** Comparison of *β*-coefficients for 205 proteins for which both protein levels at baseline and changes between baseline and first follow-up were associated with T2D onset. Pearson correlation coefficient for these two *β*-coefficients was 0.76.

**Supplementary Figure 10.**
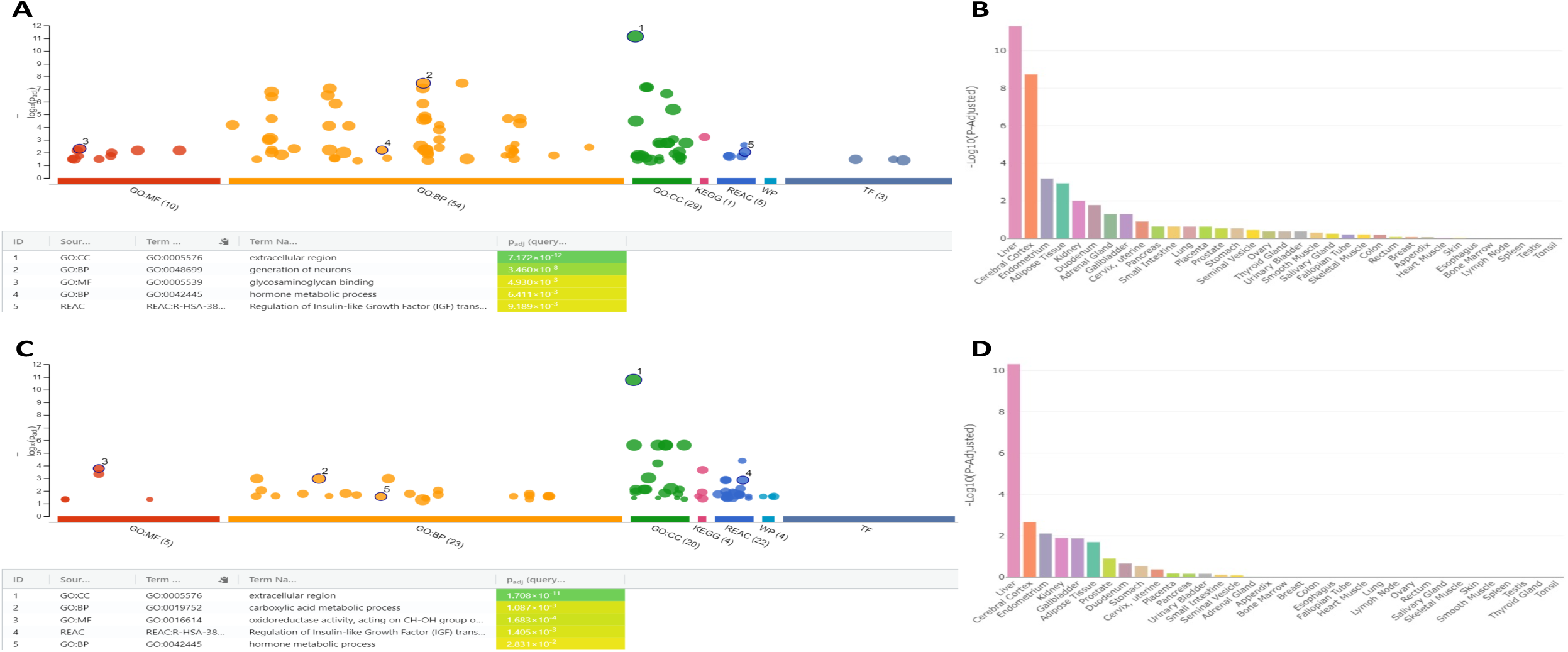
Functional enrichment analysis results for **(A – B)** 522 proteins associated with incident type 2 diabetes, and **(C – D)** subset of 205 proteins associated with longitudinal changes. Functional annotations are plotted in (**A**) and (**C**), and tissue-specific gene expression enrichment are plotted in (**B**) and (**D**).

**Supplementary Figure 11.**
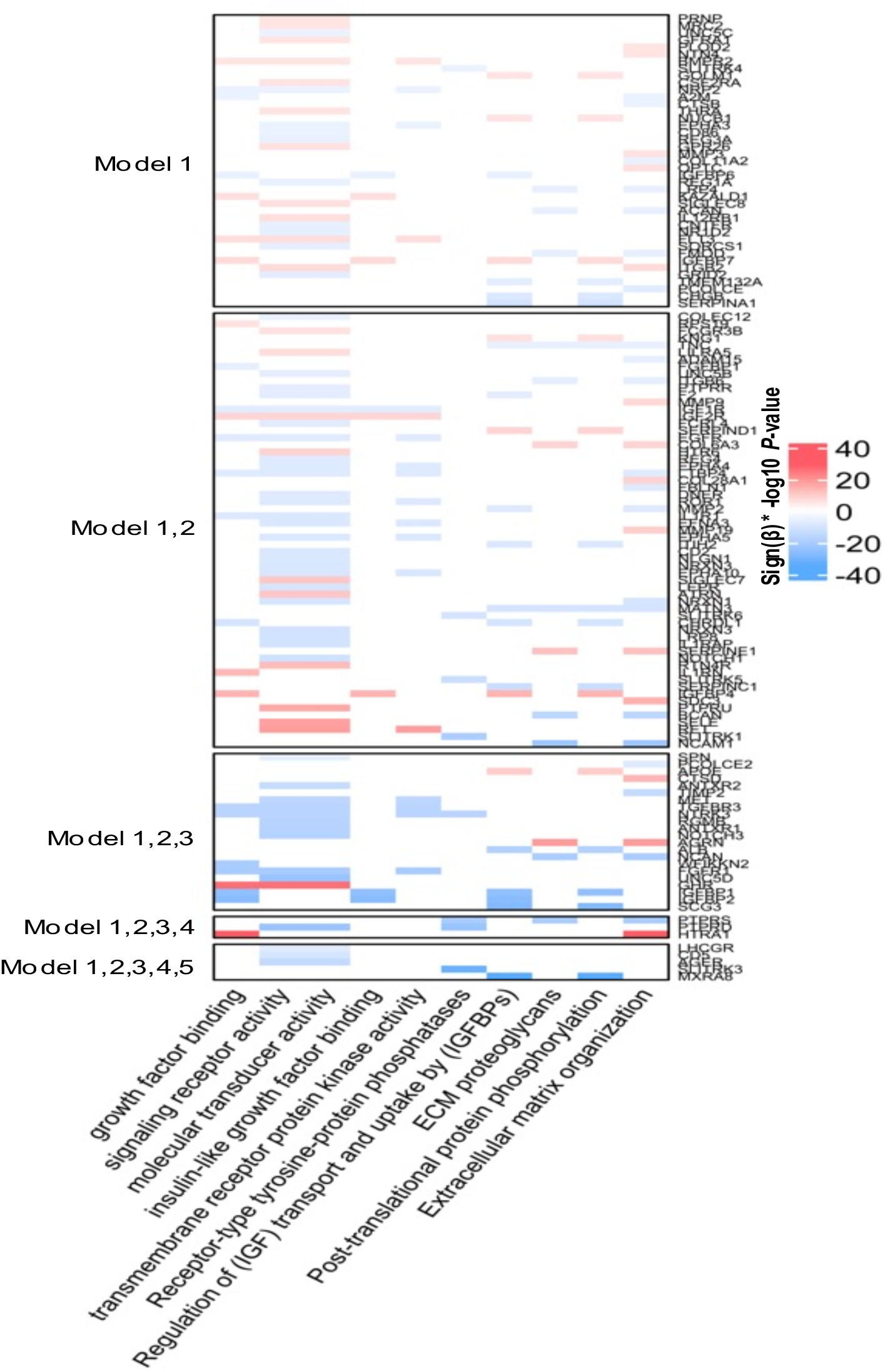
Heatmap showing proteins with significant clustering within Gene Ontology term of Molecular Function (GO:MF) and Reactome (REAC) pathways. The colors represent -log10(*P*) for protein associations with incident type 2 diabetes (T2D) in Model 1 at baseline, adjusted for age, sex, and ethnicity. Red color scale indicates positive effect size, while blue color indicates negative effect size. The x-axis plots the five significant functional enrichment pathways in GO:MF (first five columns from the left) and REAC (last five columns from the right). The 522 associated proteins from Model 1 were clustered according to their significance across Models 2 to 5. For example, ‘Model 1’ includes proteins significant only in Model 1. ‘Model 1, 2, 3, 4, 5’ indicates proteins that remained significant in all five models after adjusting for the five key T2D risk factors. Within each subgroup, proteins are ordered by their *P*-values according to the association analysis from Model 1.

**Supplementary Figure 12.**
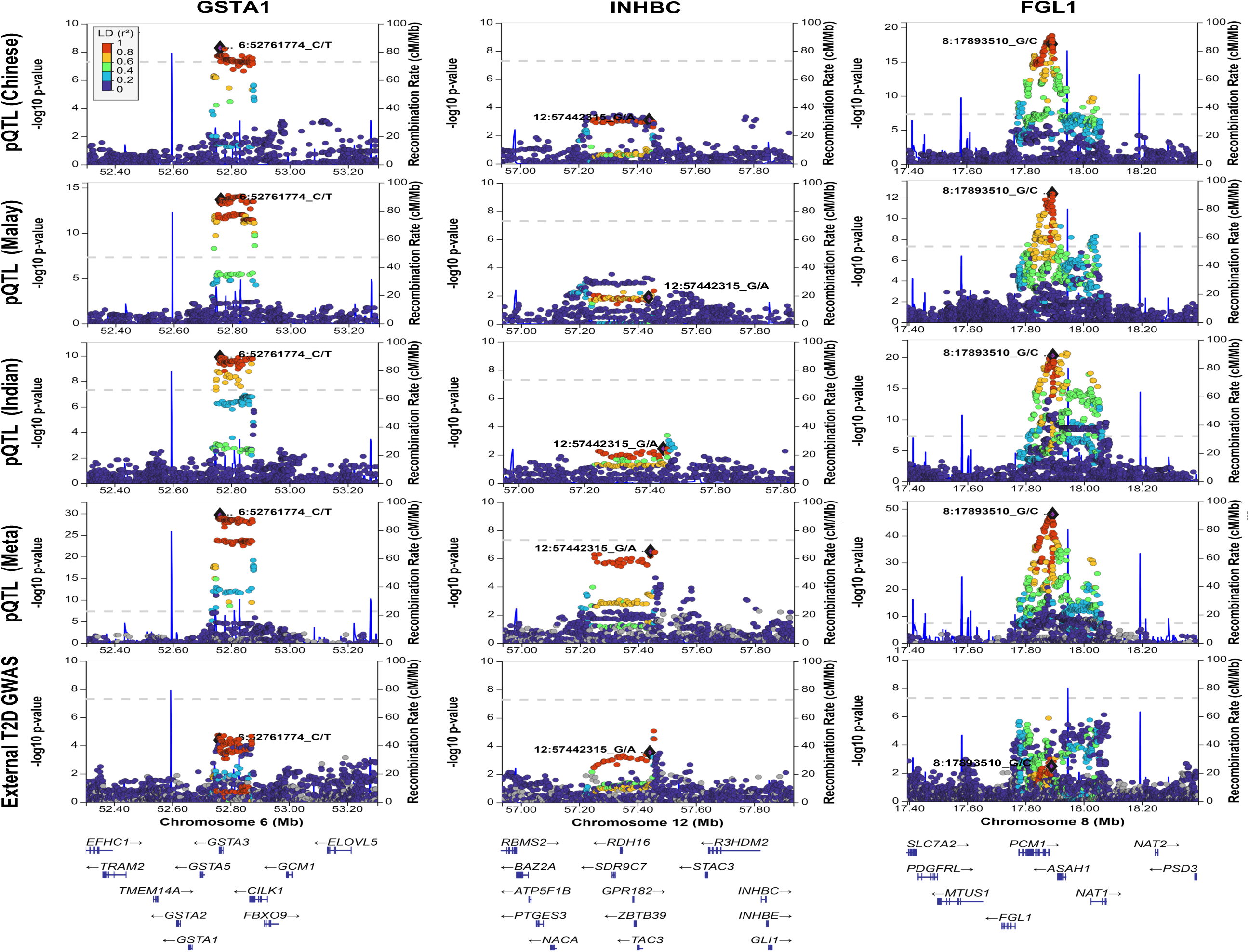
Regional association plots of genetic variants with protein levels (GSTA1, INHBC and FGL1) or type 2 diabetes (T2D) spanning ±500 kb around lead variants. The first three rows display protein quantitative trait locus (pQTL) association results in each population (Chinese, Malay, and Indian), and the fourth row shows the multi-ancestry meta-analysis pQTL association results. The last row illustrates genetic variants associated with T2D from independent multi-ancestry genome-wide association study (GWAS) (PMID: 35551307). The labeled variants, marked in purple, are the lead variant in meta-analysis of each protein: rs2749010 (6: 52761774_C/T) for GSTA1 (left column); rs2943692 (12: 57442315_G/A) for INHBC (middle column); rs11778664 (8: 17893510_G/C) for FGL1 (right column). Grey dashed line represents genome-wide significance threshold (*P* = 5 × 10^−8^). Variants are colored based on MEC1 population-specific linkage disequilibrium for population-specific association analysis (rows 1 to 3), and MEC1 Chinese linkage disequilibrium for multi-ancestry meta-analysis and T2D association. Genes in the region are displayed below the plots. Mb:megabase; cM:centimorgan

