## Supplemental Note for "Circulating proteomic profiles are associated with the onset of type 2 diabetes in a multi-ethnic Asian population – a longitudinal study"

**Supplementary Note**

1. Physical examination of participants in MEC cohort
   At each of the timepoints, waist circumference was measured at the midpoint between the last rib and iliac crest and hip circumference was measured at the greater trochanter of the femur. Systolic and diastolic blood pressures were measured twice after participants rested for 5 min. A third reading was performed if the difference between the first two readings was greater than 10 mmHg or for systolic blood pressure and greater than 5 mmHg for diastolic blood pressure. The physical examination also collected blood for blood biomarkers measurements, including insulin and triglycerides, and stored for future research [1].

2. Sample exclusion for incident type 2 diabetes (T2D) and longitudinal change analyses
   For analysis of proteomics measured at baseline and incident T2D, we excluded participants with diabetes or a history of cancer, stroke, or heart diseases at baseline (N = 23), duplicate samples (N = 4), samples that did not pass SomaLogic quality control (N = 82), missing fasting glucose information (N = 161), and missing waist circumference (WC) measurements (N = 3).
   For longitudinal analysis, to avoid reverse causation on protein level due to potential medical treatment between baseline and follow-up, we excluded diabetes cases with a self-reported diagnosis of diabetes at the follow-up visit (N = 61). We further excluded one individual with missing WC data at follow-up (**Supplementary Figure 1**).

3. Quality control processes used by SomaLogic
   The SomaScan assay was carried out in a 96-well plate, including 5 Calibrator Controls, 3 Quality Controls and 12 hybridization controls in all wells for data standardization. To correct for within-plate variability, normalization with hybridization controls was first used to remove hybridization variation, and median normalization across calibrator controls was then used to correct for within-dilution bins (20%, 0.5%, and 0.005%) variation. To correct for between-plate variability, plate scaling and calibration were performed on a per-plate basis to remove overall intensity differences between runs with Calibrator Controls. Finally, median normalization to a reference was performed on the individual samples with Quality Controls samples. During these standardization steps, multiple scaling factors were generated for each sample/aptamer at each step.

4. Comparison of association results from conditional and unconditional logistic regression
   We compared conditional logistic regression (using matched 513 cases and 789 controls pairs) with unconditional logistic regression (maximizing all available samples) when assessing protein associations with incident T2D. Both models yielded similar outcomes with the same sample set (**Supplementary Figure 5A**). However, due to sample loss during shipment, opting for unconditional analysis allowed us to utilize more samples for the analysis, thereby increasing the statistical power of the results (**Supplementary Figure 5B**).

5. Enrichment of the SomaScan protein panel and permutation analysis
   To assess potential biases caused by duplicated Entrez gene IDs of the protein compounds in SomaScan, we compared the enrichment results between with and without the removal of duplicate proteins. The enrichment results remained consistent with a perfect correlation ($r^2$ = 1.00) of the -$\log_{10}$(*P*-value). We presented results without removal of duplicates from the 4,775-protein background.
   The top enrichment of the 4,775 proteins indicated significant enrichment in the liver ($P < 1x10^{-17}$), with 185 (3.9%) proteins specifically enriched in this tissue. Of the 522 incident-

T2D associated proteins, 57 (30.8%) overlapped with the 185 liver-enriched proteins. To validate the significance of liver enrichment in our study, we conducted a permutation analysis 100 times, randomly selecting 522 proteins from the 4,775 set. Liver enrichment occurred only three times, with none of these instances reaching statistical significance ($P < 0.05$).

6. <u>Protein quantitative trait locus (pQTL) analysis and genetic instruments of proteins</u>

We identified genetic instruments by examining genetic variants within a 1Mb cis region around each protein-encoding gene. To account for different linkage disequilibrium (LD) patterns across the three ancestry groups, we conducted pQTL analysis between all genetic variants with a minor allele count greater than 5 and all 4,775 proteins separately in each ancestry group. For each ancestry, the log-transformed protein levels were adjusted using linear regression with age, sex, and sample ascertainment. Residuals from this linear regression were then rank-inverse normalized and used as phenotypes for pQTL association testing. Association analyses were conducted using the genome-wide linear regression test implemented in regenie version 3.1.3, with separate analyzes performed for each population dataset [2]. Regenie uses a two-stage approach: first, a linear mixed model adjusts for population structure; second, a sparse linear regression model tests for association. In the first stage, genetic variants included for analysis had a minor allele frequency greater than 5%, a genotyping rate above 90%, a Hardy-Weinberg equilibrium test *P*-value greater than $10^{-6}$, and were subjected to linkage disequilibrium (LD) pruning (200 variant windows, 50 variant sliding windows, and LD $r^2 < 0.2$). For each trait, leave-one-chromosome-out predictors derived from this step were utilized as covariates in the subsequent step 2 analysis. In the second stage, linear regression analysis tested the association between the residualized phenotype and the genetic marker. The association model further adjusted for the first ten principal components of ancestry. We aggregated ancestry-specific pQTL summary statistics using sample size weighted fixed-effects meta-analysis and double genomic control correction implemented in METAL (version released on 2011-03-25) [3]. Genetic instruments were identified based on locus-wide significance ($P < 0.05/n$ SNPs in the locus) and consistent directional associations across all three ancestry groups. We further refined the selection by identifying same conditionally independent signals within each locus across the three ancestry groups for further analysis. We excluded all palindromic SNPs using the 'action=3' command during data harmonization within the “TwoSampleMR” R package for Mendelian randomization analysis. To assess potential weak instrument bias in Mendelian randomization, we computed F-statistics for all the genetic instruments. The average F-statistic was 124.3 (range: 9.0 – 1098.2). Only one genetic variant (rs4888360) had an F-statistic <10 in its association with the CTRB2 protein.

<u>References</u>

1. Tan, K.H.X., et al., *Cohort Profile: The Singapore Multi-Ethnic Cohort (MEC) study.* Int J Epidemiol, 2018. **47**(3): p. 699-699j.
2. Mbatchou, J., et al., *Computationally efficient whole-genome regression for quantitative and binary traits.* Nat Genet, 2021. **53**(7): p. 1097-1103.
3. Willer, C.J., Y. Li, and G.R. Abecasis, *METAL: fast and efficient meta-analysis of genomewide association scans.* Bioinformatics, 2010. **26**(17): p. 2190-1.
